# An analysis of studies pertaining to masks in Morbidity and Mortality Weekly Report: Characteristics and quality of all studies from 1978 to 2023

**DOI:** 10.1101/2023.07.07.23292338

**Authors:** Tracy Beth Høeg, Alyson Haslam, Vinay Prasad

**Author notes:** Corresponding author: Tracy Beth Høeg, MD, PhD Department of Epidemiology and Biostatistics UCSF Mission Bay Campus | Mission Hall: Global Health & Clinical Sciences Building | 550 16th St, 2nd Fl, San Francisco, CA 94158. **Disclosure:** Vinay Prasad’s Disclosures. (Research funding) Arnold Ventures (Royalties) Johns Hopkins Press, Medscape, and MedPage (Honoraria) Grand Rounds/lectures from universities, medical centers, non-profits, and professional societies. (Consulting) UnitedHealthcare and OptumRX. (Other) Plenary Session podcast has Patreon backers, YouTube, and Substack. Tracy Beth Høeg’s Disclosures. (Honoraria) Brownstone Institute, Global Liberty Institute. (Consulting) Florida Department of Health (Other) Substack, payment for writing at The Atlantic, Los Angeles Times, New York Times, The Hill and Tablet Magazine. Alyson Haslam has no disclosures to report. **Funding:** None.

## Abstract

**Importance:** Because the MMWR has substantial influence on United States public health policy and is not externally peer-reviewed, it is critical to understand the scientific process within the journal. Mask policies during COVID is one topic that has been highly influenced by data published in the MMWR.

**Objective:** To describe and evaluate the nature and methodology of the reports and appropriateness of conclusions in MMWR pertaining to masks.

**Design, Setting and Participants:** Retrospective cross-sectional study of MMWR publications pertaining to masks from 1978 to 2023.

**Main outcome measures:** Study date, design, disease focus, setting, population and location. Whether the study was able to assess mask effectiveness, if results were statistically significant, if masks were concluded to be effective, if randomized evidence and/or conflicting data was mentioned or cited, if causal statements were made about effectiveness, and, if so, whether they were appropriate.

**Results:** 77 studies, all published after 2019, met our inclusion criteria. 75/77 (97.4%) studies were from the United States alone. All geographic regions and age groups were represented. The most common study design was observational without a comparator group 22/77 (28.6%). The most common setting was the community (35/77;45.5%). 0/77 were randomized studies. 23/77 (29.9%) assessed mask effectiveness, with 11/77 (14.3%) being statistically significant, but 58/77 (75.3%) stated masks were effective. Of these, 41/58 (70.7%) used causal language. Only one mannequin study used causal language appropriately (1.3%). 72/77 (93.5%) pertained to SARS-CoV-2 alone. None cited randomized data. 1/77 (1.3%) cited conflicting evidence.

**Conclusions and Relevance:** MMWR publications pertaining to masks drew positive conclusions about mask effectiveness over 75% of the time despite only 30% testing masks and <15% having statistically significant results. No studies were randomized, yet over half drew causal conclusions. The level of evidence generated was low and the conclusions drawn were most often unsupported by the data. Our findings raise concern about the reliability of the journal for informing health policy.

## Introduction

Prior to the COVID-19 pandemic, pooled randomized data^1^ on surgical and N95 respirator masks in the community and healthcare setting failed to demonstrate evidence of efficacy against influenza or influenza-like illness. In March of 2020, the Centers for Disease Control and Prevention (CDC) did not generally recommend mask wearing for healthy people^2^, consistent with the advice from the US Surgeon General.^3^ Over several weeks in March and early April 2020, a coordinated social media campaign to recommend masks began.^4^ Then on April 3rd, 2020, the CDC recommended people ages 2 years and older wear a cloth face covering in public.^5^ On July 15th, 2020, the CDC Director recommended all Americans start wearing masks as a way to “get the epidemic under control” ^5, 6^ citing a Morbidity and Mortality Weekly (MMWR) study involving two hairstylists in Missouri. ^7^ That coming Fall of 2020, universal masking in schools and daycares was recommended by the CDC^8^ and widespread mandates were enacted at the state, district and county levels for children as young as two. Masking on public transportation was required by federal mandate starting January of 2021.^9^

MMWR is a weekly scientific journal without external peer review overseen by the CDC to publish data on nationally notifiable infectious diseases, which can then be used for program planning, evaluation, and policy development.^10^ It is considered their primary avenue for disseminating scientific information and is often referred to as “the voice of CDC.” ^10^ The review and publication process at this journal, the levels of evidence generated, and the extent to which the studies published in this journal represent and advance current international scientific understanding remain largely opaque to the general public.

The aim of the present study was to evaluate all studies published in MMWR pertaining to masks, looking specifically at what conclusions were drawn about mask effectiveness and whether or not the conclusions were appropriate given the data presented. If causality was inferred, we determined whether or not this was appropriate, given the study’s methodology. Secondary aims included describing multiple study characteristics, including study type, number of authors, if some or all of the authors were from the CDC, and whether or not studies cited randomized or conflicting data.

## Methods

### Study identification and data abstraction

We sought to assess MMWR face mask studies by searching PubMed using two search strategies: 1) (“MMWR. Morbidity and mortality weekly report”[Journal]) AND (“face covering”) AND (covid); 2) (ℌMMWR. Morbidity and mortality weekly report”[Journal]) AND (mask) AND (covid). The searches were done on June 8, 2023. For the initial search, we included all studies, regardless of study design or publication date, and we did not have any restriction criteria in the search. After reviewing articles, we removed guidance documents and an article that was a figure only (e.g., no methods).

From each study, we abstracted the study design, setting, general age of participants (child, adolescents, adults, and/or older adults), number of people in the analysis, number of study authors, if there was a control or comparator group (yes or no) and if yes, how many were masked vs unmasked, the geographic region, whether the study tested masks, whether there was a conclusion made about mask effectiveness, if causal language was used, and if yes, was used appropriately (methodology permitted causal inference); if the conclusions matched with the study findings (i.e., the conclusions about masks were supported by the results); whether the study pointed to other evidence of mask efficacy/effectiveness and the source of evidence; if randomized data on masking was cited; and if conflicting data was cited or mentioned. Information was also obtained on the number of authors per study and whether or not any of the authors were affiliated with the CDC.

We defined geographic region of the US as such: Far West (WA, OR, CA, NV, AK, HI), Rocky Mountain (MT, ID, WY, UT, CO), Plains (ND, SD, MN, IA, NE, KS, MO), Southwest (AZ, NM, OK, TX), Great Lakes (WI, MI, IL, IN, OH), Southeast (AR, LA, MS, AL, GA, FL, KY, TN, WV, VA, NC, SC), Mideast (NY, PA, NJ, MD, DE), and New England (VT, NH, MA, CT, ME, RI).

Multicomponent mitigation strategy studies were considered as testing masks, if masks were specifically identified as being one of the components. We coded each study as testing masks or not if it included any type of control or comparison group or time period. We coded study results as being indeterminate for studies that did not test masking, no difference/negative if masking was no better, and positive if numbers were more favorable for masking, even if there were no formal statistical tests conducted. We then determined whether or not the studies testing masks had statistically significant results.

We coded the study’s conclusions about masking, according to the authors’ conclusion statements at the end of the abstract/discussion as favorable for masking or neutral (no difference). This coding was done independently by two people (AH and TBH). Causal language was defined as using terms such as, “can”, “likely”, “led to” or otherwise drawing definitive conclusions about mask effectiveness which was not based on references to other studies. We defined “appropriate” use of causal language as those that had a randomized design or observational methodology, which permitted causal inference.

### Statistical analysis

We presented descriptive characteristics and compared frequencies of study characteristics between studies testing mask efficacy or effectiveness and those that do not test mask efficacy or effectiveness. We used Chi-square and Wilcoxon rank sum test to determine differences between groups. We conducted all statistical analysis in R statistical software (version 4.6.1). Using package ‘irr’, we calculated a kappa statistic to measure the amount of agreement in whether the study determined a mask to be effective or not (including not determined). We also calculated a kappa statistic to determine whether the study authors used causal language in describing their results.

In accordance with 45 CFR §46.102(f), this study was not submitted for University of California, San Francisco institutional review board approval because it involved publicly available data and did not involve individual patient data.

## Results

Our search, spanning the years 1978 to 2023, identified 83 MMWR published studies on PubMed, all of which were published after 2019. We excluded 5 guidance documents and a search result that was a stand-alone figure. Our search identified 83 MMWR published studies on PubMed. We excluded 5 guidance documents and a search result that was a stand-alone figure. Of the included 77 studies, 23 (29.9%) studies were graded as assessing mask effectiveness with the remaining 54 (70.1%) not having the methodology to do so. 72 (93.5%) pertained to SARS-CoV-2, one pertained to SARS-CoV-2 and influenza co-infection, three studies pertained mainly to influenza and one pertained to rhino and enteroviruses.

No studies met our inclusion criteria from 1978 through 2019. Thirty studies were published in 2020; 33 were published in 2021; and 14 were published in 2022. The median number of participants was 558 (IQR: 171, 2964). The median number of authors was 13 (IQR: 9, 26), total listed authors including duplicates was 1544. Seventy studies (90.9%) had one or more authors affiliated with the CDC.

The kappa statistic for intra-author agreement in the determination of whether studies made a conclusion about masks was 0.69 (p<0.0001), and the kappa statistic for the use of causal language was 0.66 (p<0.0001). These numbers suggest that the agreement was substantial for both.

Study characteristics, stratified by whether or not masks were tested for effectiveness, are shown in Table 1 and described in detail in the supplementary material. 75/77 (97.4%) studies were from the United States alone, one was from Chile and one was from multiple countries. All geographic regions were represented with 32/77 (41.6%) using multi-state data. The most common study design was observational without a control or comparator group 22/77 (28.6%). All age groups were represented. The most common setting was community (35/77; 45.5%) followed by kindergarten through high school (13/77; 16.9%). The characteristics of the 77 studies, by whether or not they had appropriate methodology to test masks, are described in the **Supplementary Material**.

**Table 1.**
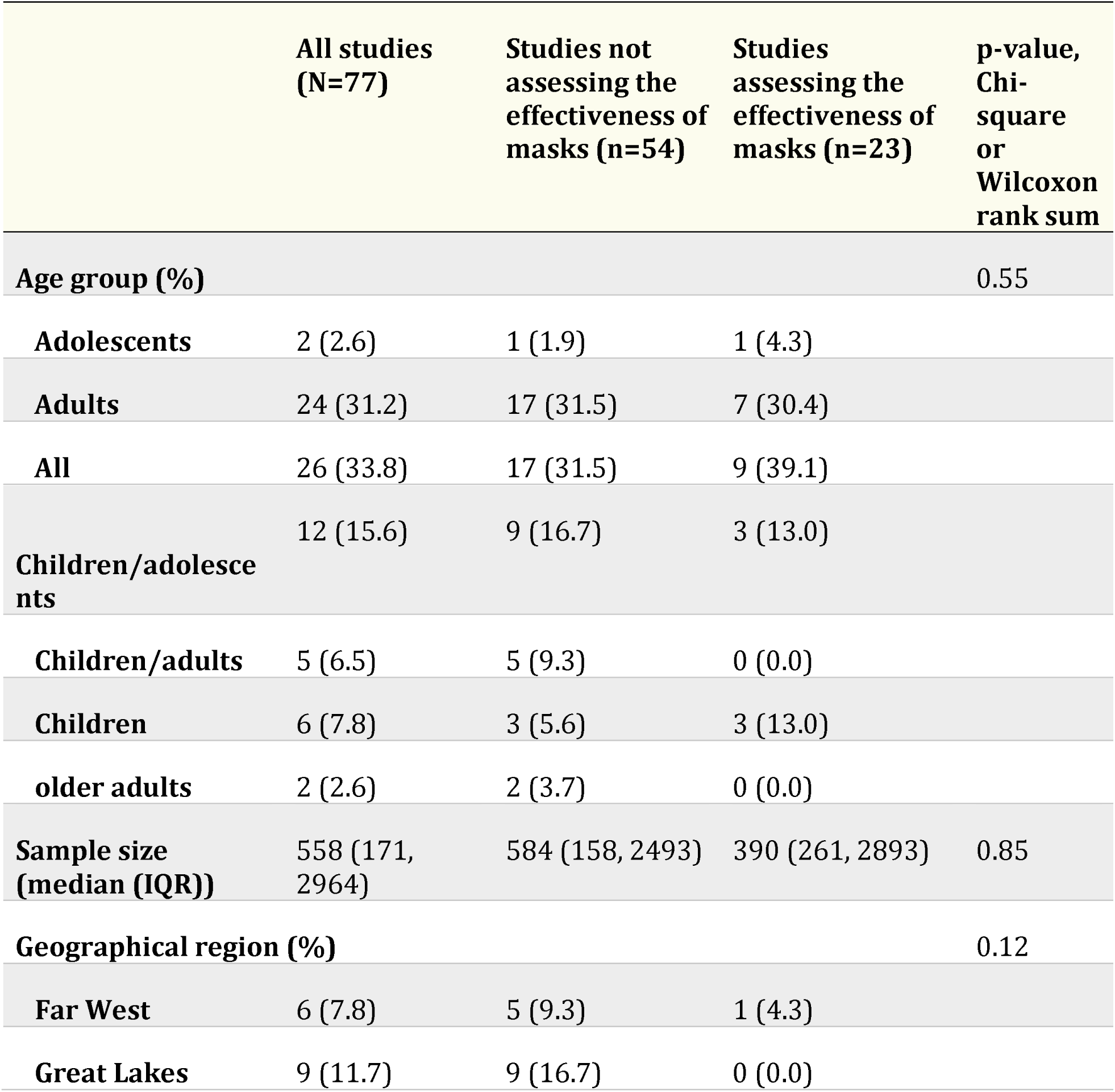
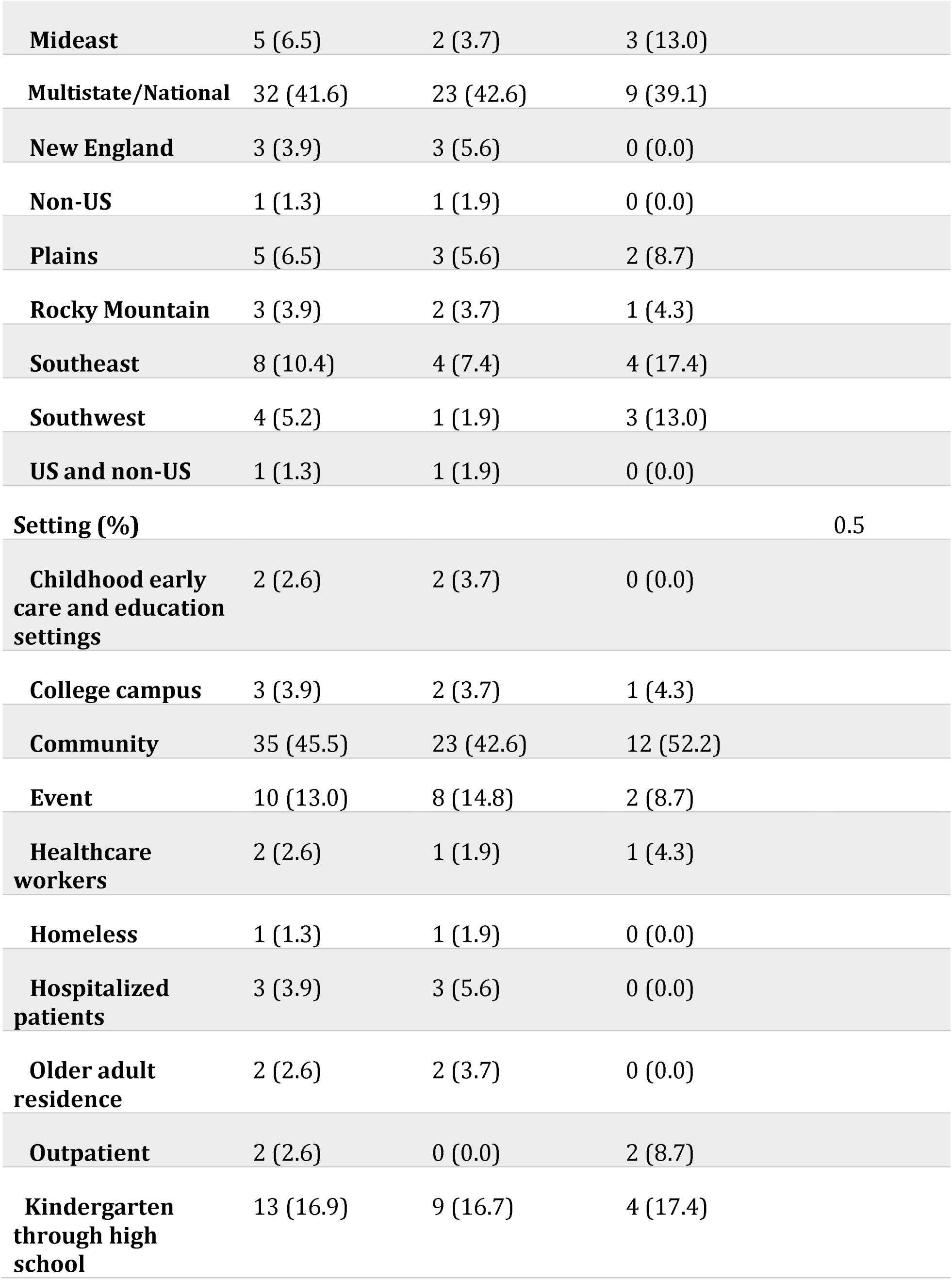
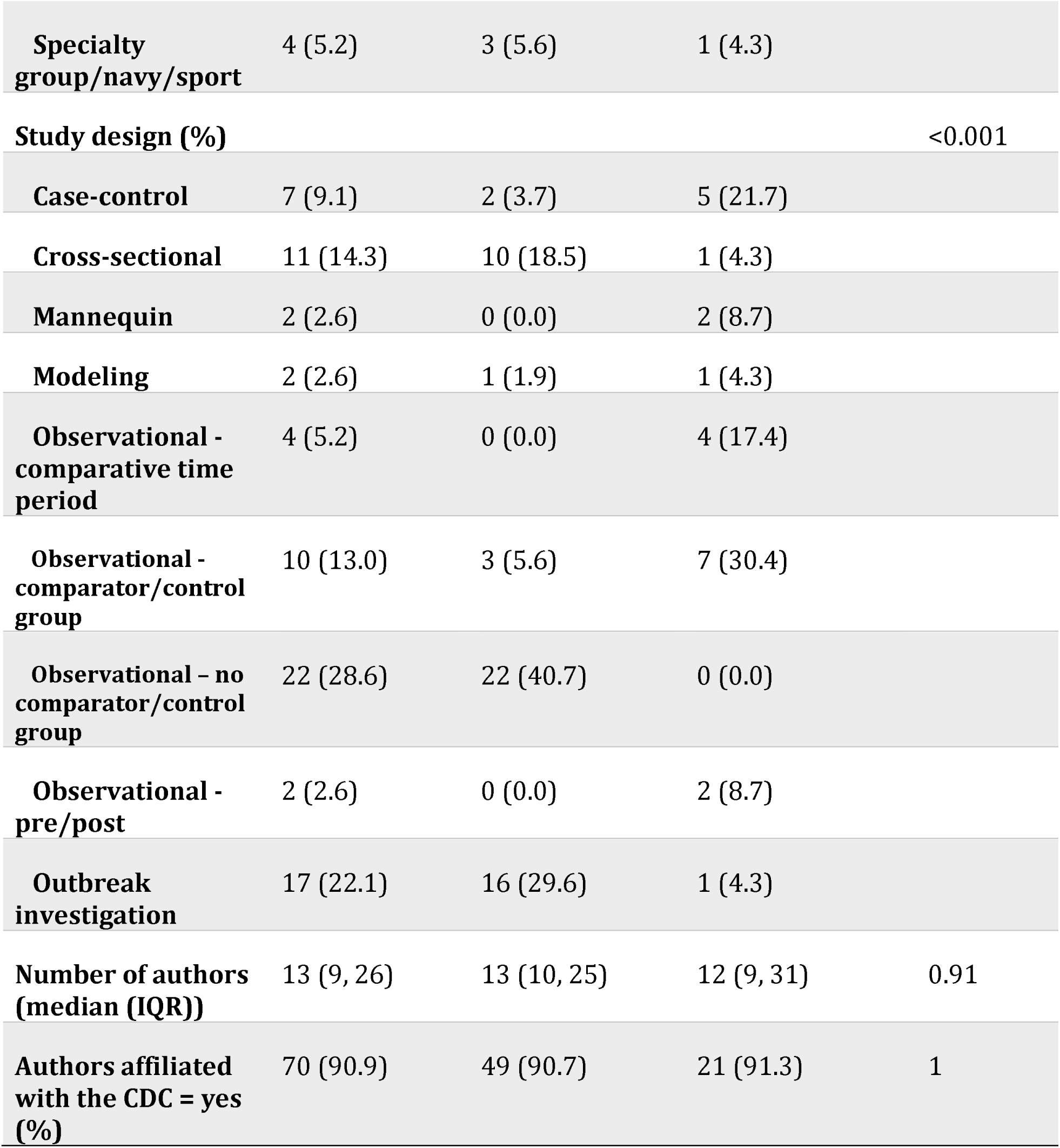
Study characteristics of Morbidity and Mortality Weekly Report studies mentioning masks or face coverings, stratified by whether mask effectiveness was tested

In Figure 1, we show a total of 23/77 (29.9%) identified studies that assessed the effectiveness of masks, however, 58/77 (75.3%) stated masks were effective. Of these 58 studies, 41/58 (70.7%) used causal language and 40/58 (69.0%) used causal language inappropriately. One mannequin study allowed causal inference. 11/77 (14.3%) found a statistically significant inverse relationship between masking and cases. No studies (0/77; 0%) were randomized. 4/77 (5.2%) had a numerically higher number of cases in the mask group than the comparator group but all 4/4 (100%) concluded masks were effective. Of all publications included, 0/77 (0.0%) cited a randomized study or review of only randomized studies. Of all 58 studies stating masks were effective, only 1/58 (1.7%), which mainly focused on influenza^11^ mentioned conflicting data on mask effectiveness.

**Figure 1.**
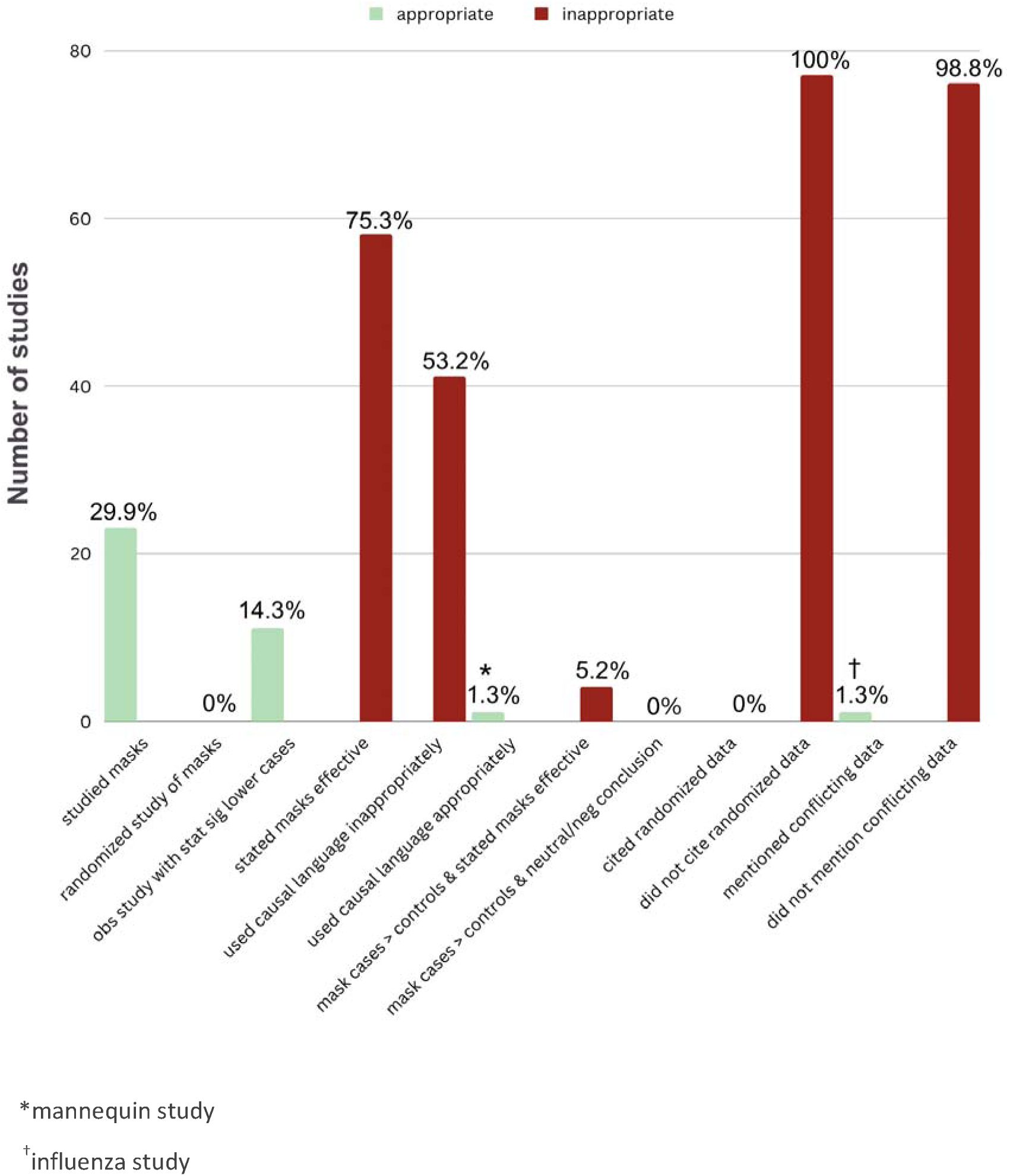
Select characteristics of the 77 Morbidity and Mortality Weekly Report (MMWR) publications pertaining to masks

Table 2. Shows examples of language used in multiple MMWR studies which did not have appropriate methodology to draw conclusions about the effectiveness of masking.

**Table 2.** Select conclusion statements indicating a causal relationship between mask wearing and decreased case rates/transmission from MMWR mask studies which failed to find evidence of mask effectiveness

| Study and date | Conclusion Statement | Control group/time period | Provided evidence of mask effectiveness |
| --- | --- | --- | --- |
| Outbreak investigation of two Missouri hairdressers 7/14/2020<sup>1</sup> | "Consistent and correct use of face coverings, when appropriate, is an important tool for minimizing spread of SARS-CoV-2" | No | No |
| Georgia camp outbreak investigation 8/7/2020<sup>2</sup> | "Consistent and correct use of cloth masks should be emphasized as important strategies for mitigating transmission" | No | No |
| Investigation of university soccer team outbreaks 10/30/2022<sup>3</sup> | "Improved strategies to promote mask use and social distancing among college-aged adults need to be implemented" | No | No |
| Hopi Tribe Outbreak investigation 11/6/2020<sup>4</sup> | "this investigation highlights the need for... encouraging consistent mask wearing" | No | No |
| Household transmission in Wisconsin and Tennessee 11/6/2020<sup>5</sup> | "all household members wearing a mask in shared spaces, can reduce the probability of household transmission." | No | No |
| Georgia school outbreak investigation 3/12/2021<sup>6</sup> | "correct mask use and physical distancing, even after educators are vaccinated, will be critical" | No | No |
| Transmission in Salt Lake City Utah Elementary Schools 3/26/2021<sup>7</sup> | "schools can be opened safely with minimal in-school transmission when critical prevention strategies including mask use are implemented" | No | No |
| Outbreaks at a youth camp and men's conference 9/3/2021<sup>8</sup> | "This investigation underscores the impact of secondary SARS-CoV-2 transmission during large events such as camps and conferences when COVID-19 prevention strategies, including vaccination, masking, physical distancing, and screening testing, are not implemented." | No | No |
| Marin County Elementary School Outbreak 9/3/2021<sup>9</sup> | "strict adherence to multiple nonpharmaceutical prevention strategies, including masking, are important to ensure safe school instruction" | No | No |
| Evaluation of a test to | "TTS strategy with multiple prevention | No | No |
| stay (TTS) strategy in Lake County, IL 12/31/2021<sup>10</sup> | components, including masking and physical distancing, resulted in low secondary transmission of SARS-CoV-2 in K–12 schools in Lake County, Illinois.” |  |  |
| Antigen test positivity after COVID-19 isolation 2/25/2022<sup>11</sup> | “The high percentage of positive antigen test results during the 5–9 days after symptom onset reinforces the importance of correct and consistent mask use during this period ” | No | No |

Table 3 shows the data characteristics of the included MMWR studies, overall and stratified by the testing of masks or not.

**Table 3.** Data characteristics of Morbidity and Mortality Weekly Report publications mentioning mask or face covering (n=77)

|  | All studies<br>(N=77) | Studies not<br>assessing the<br>effectiveness of<br>masks (n=54) | Studies<br>assessing the<br>effectiveness of<br>masks (n=23) | p-value,<br>Chi-square |
| --- | --- | --- | --- | --- |
| **Conclusion**<br>**supportive of masks =**<br>**yes (%)** | 58 (75.3) | 36 (66.7) | 23 (100.0) | 0.016 |
| **Results (%)** |  |  |  | <0.001 |
| Positive | 19 (24.7) | 1 (1.9) | 18 (78.3) |  |
| Indeterminate | 54 (70.1) | 53 (98.1) | 1 (4.3) |  |
| No difference | 4 (5.2) | 0 (0.0) | 4 (17.4) |  |
| **Tested for statistical difference (%)** |  |  |  | <0.001 |
| Yes | 15 (19.5) | 0 (0.0) | 15 (65.2) |  |
| No | 10 (13.0) | 2 (3.7) | 8 (34.8) |  |
| Not applicable | 52 (67.5) | 52 (96.3) | 0 (0.0) |  |
| **Use of causal language (%)** |  |  |  | 0.009 |
| Yes | 41 (53.2) | 25 (46.3) | 16 (69.6) |  |
| No | 19 (24.7) | 12 (22.2) | 7 (30.4) |  |
| Not applicable | 17 (22.1) | 17 (31.5) | 0 (0.0) |  |
| **Appropriate use of causal language (%)** |  |  |  | 0.011 |
| Yes | 1 (1.3) | 0 (0.0) | 1 (4.3) |  |
| No | 40 (51.9) | 25 (46.3) | 15 (65.2) |  |
| Not used | 19 (24.7) | 12 (22.2) | 7 (30.4) |  |
| Not applicable | 17 (22.1) | 17 (31.5) | 0 (0.0) |  |
| **Cited other mask** | 40 (51.9) | 23 (42.6) | 17 (73.9) | 0.023 |
| **effectiveness studies**<br>**= yes (%)** |  |  |  |  |
| **Cited randomized**<br>**mask data = yes (%)** | 1 (1.3) | 1 (1.9) | 0 (0.0) | 1 |
| **Cited conflicting mask**<br>**data = yes (%)** | 1 (1.3) | 1 (1.9) | 0 (0.0) | 1 |

As shown in Table 4, of the studies that evaluated masks, 22/22 (100%) concluded masks were effective; 18/22 (81.8%) reported results favoring masks; 13/22 (59.1%) tested for statistical differences; and 12/22 (54.5%) of which were statistically significant.

**Table 4.**
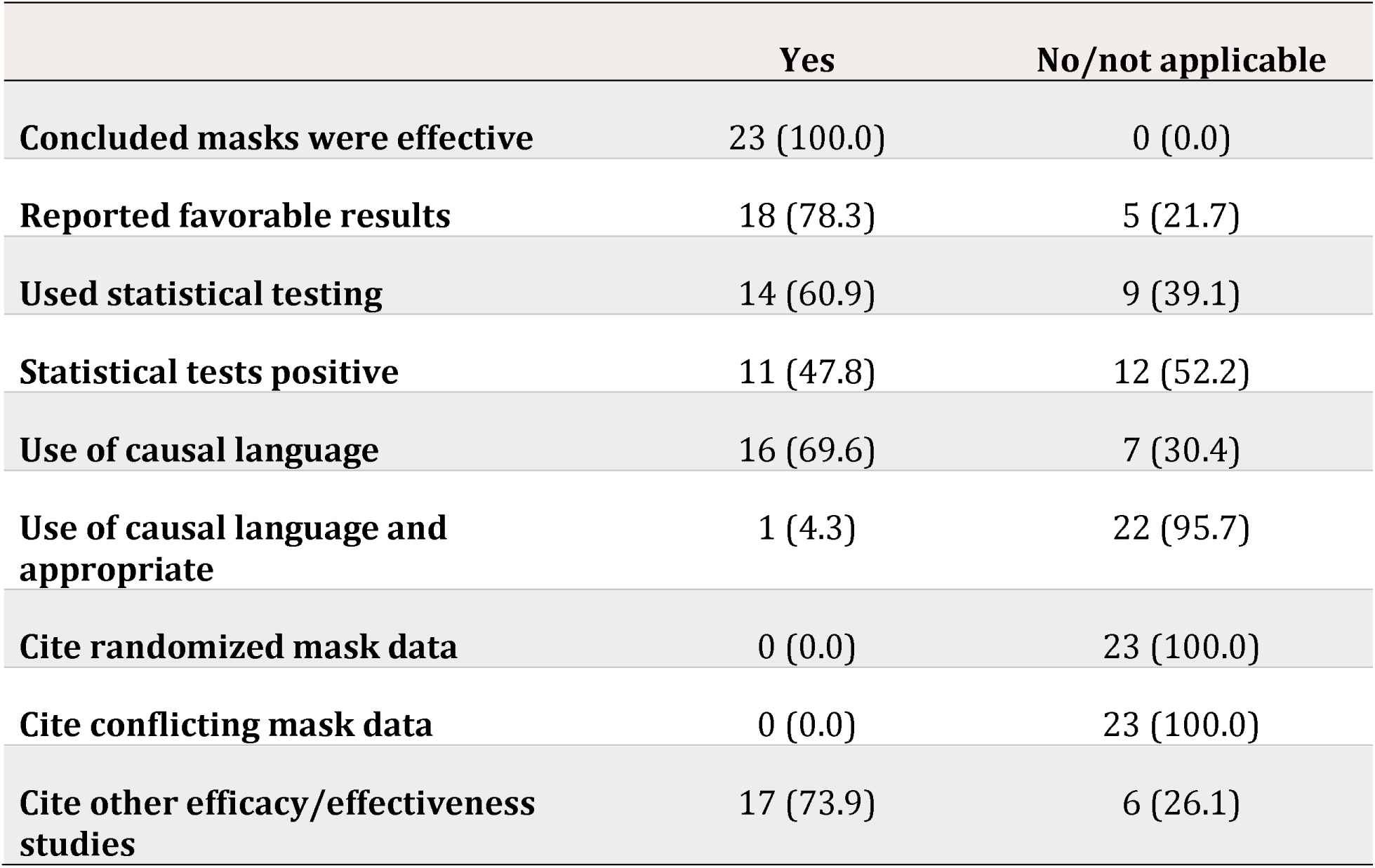
Characteristics of Morbidity and Mortality Weekly Report publications evaluating studies testing masks (n= 23)

|  | **Yes** | **No/not applicable** |
| --- | --- | --- |
| **Concluded masks were effective** | 23 (100.0) | 0 (0.0) |
| **Reported favorable results** | 18 (78.3) | 5 (21.7) |
| **Used statistical testing** | 14 (60.9) | 9 (39.1) |
| **Statistical tests positive** | 11 (47.8) | 12 (52.2) |
| **Use of causal language** | 16 (69.6) | 7 (30.4) |
| **Use of causal language and**<br>**appropriate** | 1 (4.3) | 22 (95.7) |
| **Cite randomized mask data** | 0 (0.0) | 23 (100.0) |
| **Cite conflicting mask data** | 0 (0.0) | 23 (100.0) |
| **Cite other efficacy/effectiveness**<br>**studies** | 17 (73.9) | 6 (26.1) |

Details about the included studies and grading of subjective endpoints are publicly available at the following GitHub repository https://github.com/tracybethhoeg/mmwrmasks under an MIT license.

## Discussion

We found that, among the 77 studies identified pertaining to masks published in MMWR, 30% tested the effectiveness of masks, with 14% having statistically significant results, yet over 75% of all 77 studies concluded masks were effective. Of the 5% that reported higher numbers of cases in the masked group than the comparator group, all concluded masks were effective. MMWR studies consistently drawing conclusions about mask effectiveness without supporting evidence is particularly problematic and difficult to justify considering the totality of randomized evidence about the use of surgical or N95 masks to prevent the spread of respiratory viruses has been negative.^1, 12^

Over 50% of the identified studies used causal language in their conclusions about mask effectiveness. Only one of these studies, which was a mannequin study, had methodology which permitted causal inference. In other words, the remaining 40 studies used language that indicated with certainty that masks lower transmission rates in spite of the fact their results, at most, found a correlation. 25 of these 40 studies, however, did not even test mask effectiveness. We have provided examples of study conclusions, which stated masks resulted in case reductions, in spite of the fact none of the studies had the appropriate methodology to assess mask effectiveness (Table 2). There were a total of 25 studies that did not evaluate masks but made causal claims about their effectiveness. It is important to note that the one identified study, which permitted causal inference, was a study of particle filtration on mannequins^13^, with unknown relevance for human health.

The inappropriate use of causal language used in MMWR studies was also adopted directly by the CDC director when she cited an observational phone survey, which also happened to be included in the present analysis^14^, stating to the public “Masks can help reduce your chance of #COVID19 infection by more than 80%.”^15^ This referenced study found an association between respondents’ recollection about mask wearing and self-reported COVID-19 tests, which was non-significant for cloth masks.

A number of studies that were particularly influential in shaping policy recommendations around masking in the public and schools were not even among the studies that attempted to properly evaluate masks, as they had no control group or comparative time period. These studies included the investigation of two Missouri hairdressers^7^, the Georgia overnight camp outbreak investigation^16^, and the Marin school outbreak investigation.^17^ None of these had methodology that permitted an evaluation of mask effectiveness, but they nonetheless drew conclusions about mask effectiveness (Table 2), which were then rapidly communicated to the public via the CDC.

Commensurate with the existing randomized data prior to 2020,^1^ the CDC had previously recommended against wearing masks to prevent respiratory infections. ^2, 6^ A shift in messaging for the public to wear masks to control the pandemic came July 15th from the CDC director. This change came following the report of two hairdressers wearing masks while working, which concluded that masks were “likely a contributing factor in preventing transmission of SARS-CoV-2 during the close-contact interactions between stylists and clients.”^6^ In this instance, public health recommendations shifted largely based on anecdotal data in MMWR.

Randomized studies are the most reliable method of determining whether an intervention is efficacious. None of the studies identified in MMWR were randomized and none cited randomized data. Due to a high likelihood of confounding variables and/or spurious findings,^18^ observational studies of masking are unlikely to provide reliable information about the ability of masks to prevent infection with or transmission of SARS-CoV-2 or other respiratory viruses and are, with few exceptions, inappropriate for causal inference.

Only one study mentioned conflicting data on masking efficacy, in spite of the existing overall negative randomized data.^1, 12^ Interestingly the focus of this study was influenza and it was an international study.^11^

Taken together, the absence of randomized data, the lack of acknowledgment of conflicting or randomized data on mask efficacy, and the tendency to conclude masks are efficacious either without any, or sufficient, data to make causal claims, is suggestive of bias within the journal. Our findings may also help explain why CDC remains an international outlier in continuing to recommend masks for COVID-19 under certain circumstances, including for children down to age 2.^19^

Concerns about publication bias within MMWR have been raised previously, when follow-up data to a 2-week study with a limited sample were submitted, which failed to identify evidence of school mask mandate effectiveness, was rejected from the MMWR.^18, 20^ In another school masking study, errors in data analysis and methodology, which normally would warrant retraction, were not addressed by the journal.^21^

One Journal of the American Medical Association (JAMA) Viewpoint^22^ described how, starting September 11, 2020, political appointees may have “demanded the ability to review and revise scientific reports” in MMWR, and concern was raised about “political appointees trying to influence the scientific process.” The extent to which this happened or is still happening is unclear. However, even prior to this, unlike other peer-reviewed scientific journals, MMWR publications have not and do not undergo any external peer review. Rather, they undergo a “clearance process”^10, 23^, which is sometimes referred to as “internal peer review”.^22^ Both political involvement and lack of input from external domain experts could influence the journal’s ability to objectively evaluate scientific data. But the extent to which either of these explain our findings is beyond the scope of this paper.

However, the process by which scientific data are interpreted and published in MMWR and then promoted by the CDC is not transparently communicated to the public. Because the CDC uses data published in the MMWR journal to develop its guidelines, the quality of scientific data and data interpretation within the journal have major implications for public health and well-being in the United States, extending far beyond masks.

Our search did not identify any mask articles published prior to 2020, though our search was not restricted by date. Ninety percent of mask studies published in the MMWR had one or more authors with CDC affiliations. There was a median of 13 authors per paper and, though there were some authors who co-authored multiple papers, there were a total of 1544 paper authors, which speaks to the large amount of effort that went into studying and publishing about this topic in the journal. It is thus disappointing that, due to the intrinsic limitation of the study designs, the sum of the work was inconclusive, yet strong conclusions were drawn and communicated to the public nonetheless.

### Limitations

Our study has important limitations. Some of the study characteristics we graded were subjective. For those (tone of conclusion, use of causal language), we used a double-blinded system and kappa statistics suggested there was substantial agreement about the categorization of studies. Second, our search criteria were broad and resulted in a number of studies that did not specifically study masks, thus our overall findings are not representative of studies of masks alone. However, this strategy did allow us to identify numerous studies which drew conclusions about masks without having a study design that could evaluate their effectiveness.

### Conclusion

We found that, while less than 20% of MMWR studies pertaining to masks generated any statistical evidence of mask effectiveness and no randomized investigations were published, more than 75% of the publications arrived at a favorable conclusion about using masks, and 70% of studies testing masks used causal language. Similarly, language about the studies’ implications, including the importance of masking, was used in multiple publications in spite of lack of supporting evidence.

None of the MMWR studies were randomized and none mentioned higher-quality randomized studies, which fail to find evidence of mask effectiveness. The extent to which our findings apply to scientific topics beyond masks is outside of the scope of our investigation. However, with regards to the topic of mask effectiveness, our findings highlight the journal’s lack of reliance on high-quality data and a tendency to make strong but unsupported causal conclusions about mask effectiveness.

## Data Availability

All data produced are available online at https://github.com/tracybethhoeg/mmwrmasks

