## Supplementary Material for "An analysis of studies pertaining to masks in Morbidity and Mortality Weekly Report: Characteristics and quality of all studies from 1978 to 2023"

*Study characteristics stratified by testing or not testing mask effectiveness*

The study characteristics, overall and stratified by mask testing or not, are presented in **Table 1**. For studies not testing masks (n=54), the age groups with the highest percentage of studies were all age groups (31.5%; n=17) and adults (31.5%; n=17). Many studies included participants from multiple regions (44.4%; n=24), but the single region with the most studies was the Great Lakes region (16.7%; n=9), followed by the Far West (9.3%; n=5). The most common setting was in the community (42.6%; n=23), followed by the kindergarten through high school setting (15.1%; n=8). The most common study design was an observational study with no comparator (including pre/post; 40.7%; n=22), followed by outbreak investigation/contact tracing (29.6% n=16). The median number of authors for these 54 studies was 13 (IQR: 9, 26).

For studies testing masks (n=23), the age groups with the highest percentage of studies were all age groups (39.1%; n=9) and adults (30.4%; n=7). Many studies included participants from multiple regions of the US (34.8%; n=8), but the single region with the most studies was the Southeast region (17.4%; n=4). The most common setting was in the community (52.2%; n=12), followed by the kindergarten through high school setting (17.4%; n=4). The most common study design was an observational study with a comparator (including pre/post; 39.1%; n=9), followed by case-control (21.7% n=5). Two (of 23; 8.7%) were mannequin studies, 1/23 (4.3%) was a modeling study, and none were randomized trials. The median number of authors on these 23 studies was 12 (IQR: 9, 31).
